# Converting between the International Prostate Symptom Score (IPSS) and the Expanded Prostate Cancer Index Composite (EPIC) Urinary Subscales: Modeling and External Validation

**DOI:** 10.1101/2023.09.20.23295834

**Authors:** Paul Windisch, Ivo Becker, Hongjian Tang, Christina Schröder, André Buchali, Daniel M. Aebersold, Daniel R. Zwahlen, Robert Förster, Mohamed Shelan

**Author notes:** Corresponding author **Correspondence:** Paul Windisch, MD, Department of Radiation Oncology, Kantonsspital Winterthur, Brauerstrasse 15, Haus R, 8400 Winterthur. Contributed equally (Robert Förster, Mohamed Shelan). **Funding:** No funding was received for this project. **Author contributions:** Conceptualization, P.W., R.F.; methodology, P.W.; formal analysis, P.W., I.B., H.T., C.S.; data curation, P.W., I.B.; writing—original draft preparation, P.W., I.B.; writing—review and editing, H.T., C.S., A.B., D.M.A., D.R.Z., R.F., M.S.; supervision, R.F., M.S.; project administration, D.R.Z.; All authors read and approved the final manuscript.

## Abstract

**Background:** The 50-item Expanded Prostate Cancer Index Composite (EPIC) and the International Prostate Symptom Score (IPSS) are two widely used options to assess prostate-related quality of life (QoL), but there is no method to convert between the two. We, therefore, developed and externally validated models for this purpose.

**Methods:** 347 consecutive patients who had previously received radiotherapy and surgery for prostate cancer at two institutions in Switzerland and Germany were contacted via mail and instructed to complete both questionnaires. The Swiss cohort was used to train and internally validate different machine learning models using 4-fold cross-validation. The German cohort was used for external validation.

**Results:** Converting between the EPIC Urinary Irritative/Obstructive subscale and the IPSS using linear regressions resulted in mean absolute errors (MAEs) of 3.88 and 6.12 below the respective previously published minimal important differences (MIDs) of 5.2 and 10 points. Converting between the EPIC Urinary Summary and the IPSS was less accurate with MAEs of 5.13 and 10.45, similar to the MIDs. More complex model architectures did not result in improved performance.

**Conclusions:** Linear regressions can be used to convert between the IPSS and the EPIC Urinary subscales. While the equations obtained in this study can be used to compare results across clinical trials, they should not be used to inform clinical decision-making in individual patients.

## Introduction

Patient-reported outcome measures (PROMs) such as quality of life (QoL) are becoming increasingly important in clinical research.^1,2^ For diseases of the prostate, such as prostate cancer (PCa) or benign prostate hyperplasia (BPH), several validated QoL questionnaires exist.^3^

While a certain degree of heterogeneity is desirable due to the different focus areas of the questionnaires, it also makes comparisons across studies difficult.^4^ This has led to trials requiring patients to complete different QoL questionnaires, sometimes at the same point in time. While this is not only cumbersome for the patient, having to answer more questions has also been shown to reduce the likelihood of a questionnaire being completed.^5^

Two common questionnaires to assess prostate-related QoL are the 50-item Expanded Prostate Cancer Index Composite (EPIC) and the International Prostate Symptom Score (IPSS).^6,7^

The EPIC consists of 50 Likert items that are used to compute four subscales: Urinary, Bowel, Sexual, and Hormonal. Each subscale has two subdomains to assess symptom severity (Function) and its effect on QoL (Bother). The Urinary subscale has two additional subdomains called Incontinence and Irritative/Obstructive. Scores range from 0 - 100, with higher scores indicating better QoL.

The IPSS consists of eight Likert items. The first seven relate to lower urinary tract symptoms of BPH, while the eighth asks about the symptoms’ effect on QoL. The first seven items are added to calculate the total, which ranges from 0 - 35, with higher scores indicating higher symptom burden.

To address the problem of converting between questionnaires, several publications have attempted to derive conversion rules.^8–10^ However, to the best of our knowledge, there is currently no established method to convert between the IPSS and EPIC. Vertosick and colleagues attempted conversions by taking only a subset of questions from QoL instruments and calculating conversion factors.^3^ However, they were unable to compare IPSS and EPIC due to differences in the domains addressed by the questionnaires. The purpose of this study was, therefore, to collect data for training as well as internally and externally validating models to enable converting between the two.

## Methods

The study was conducted in radiation oncology departments at two institutions, the Cantonal Hospital Winterthur in Switzerland and the Ruppiner Kliniken GmbH in Germany.

Three hundred and forty-seven consecutive patients who had received radiation therapy for prostate cancer in the post-operative setting at our institutions between 2010 and 2020 were identified and received the German versions of the EPIC and IPSS questionnaires in August 2020, unless a date of death had been documented in our electronic health records.

We received responses from 208 patients. Of these responses, 175 had no missing values for any of the quality of life questionnaires and a signed informed consent.

Training and internal validation were performed on the Swiss cohort (n = 142) using cross validation, while the German cohort was stored for external validation (n = 33) to assess if the model generalizes well to previously unseen data from another institution without showing signs of overfitting.

We used a three-step approach: First, we visualized relevant patient characteristics to ensure that there was a correlation between the EPIC and IPSS scores, that patient characteristics were similar in both the training and external validation set, and that both sets contained a variety of different scores from bad over mediocre to excellent.

Second, we developed four baseline models that all had one input each: A model to predict the EPIC Urinary Summary score when only the total IPSS is known, a model to predict the IPSS when only the EPIC Urinary Summary is known, a model to predict the Epic Urinary Irritative/Obstructive score when only the total IPSS is known and a model to predict the total IPSS when only the EPIC Urinary Irritative/Obstructive score is known. For all baseline models, we used a simple linear regression.

In the third step, we tried to improve upon the performance of the baseline models by using more complex machine learning algorithms and using the raw answers to the questionnaires instead of the computed scores. For the purpose of this article, we use the term advanced models as a reference to models trained in this step. For every task, we used a linear regression, a support vector regression, a k-nearest neighbors regression, and an XGBoost, respectively.^11^ In turn, we trained four models each for the following tasks: Predicting the EPIC Urinary Summary score using all IPSS questions. Predicting the EPIC Urinary Irritative/Obstructive score using all IPSS questions. Predicting the total IPSS using all EPIC questions that are used for the computation of the EPIC Urinary subscale. Predicting the total IPSS using only the most relevant EPIC questions that are used for the computation of the EPIC Urinary subscale.

Questions were considered relevant if the authors considered the content of the question to be reflected in one or multiple questions of the IPSS. We selected questions 6d, e, and f, which ask about weak urine stream or incomplete emptying, waking up to urinate, and the need to urinate frequently during the day.

All models were trained and internally validated using 4-fold cross-validation, and the mean absolute error (MAE) was used for scoring. Hyperparameter tuning was performed using a randomized search with 250 iterations each, and the respective ranges can be found in the code (see below).^12^

Data preprocessing, analysis, and visualization were performed with Python (version 3.9.7) using the numpy (version 1.20.3), pandas (version 1.3.4), scikit-learn (version 0.24.2), matplotlib (3.4.3), and seaborn (0.11.2) packages.

The full dataset, notebook, environment file, and trained models have been uploaded to a public repository (https://github.com/windisch-paul/EPIC-IPSS-converter).

Some of the patients in the dataset have also been analyzed in another publication on the correlation between dose-volume histogram parameters and quality of life in patients with prostate cancer treated with surgery and radiotherapy.^13^

Institutional review board approval was obtained from the ethical review committee of the canton of Zurich for a project (project number: BASEC 2020-02112) to analyze the effects and side effects of radiotherapy at our institution (ClinicalTrials.gov Identifier: NCT05192876). Written informed consent for the analysis of anonymized clinical and imaging data was obtained from all patients, and all data were gathered in accordance with the World Medical Association Declaration of Helsinki: Research involving human subjects.

### Data availability

All data and code used to obtain the results of this study have been uploaded to https://github.com/windisch-paul/EPIC-IPSS-converter.

## Results

Selected patient characteristics and their distributions are visualized in Figure 1. The median age at the survey was 72.5 years for the training set (range: 53.5 - 85.7 years) and 69.2 years for the external validation set (range: 53.2 - 82.7 years). The median IPSS was 6 for the training set (range: 0 - 28) and 7 for the external validation set (range 2 - 32). The median EPIC Urinary Summary score was 82 for the training set (range: 22.9 - 100) and 73.6 for the external validation set (range: 37.5 - 82.7). The median EPIC Urinary Irritative/Obstructive score was 89.3 for the training set (range: 35.7 - 100) and 82.1 for the external validation set (range: 39.3 - 92.9).

**Figure 1.**
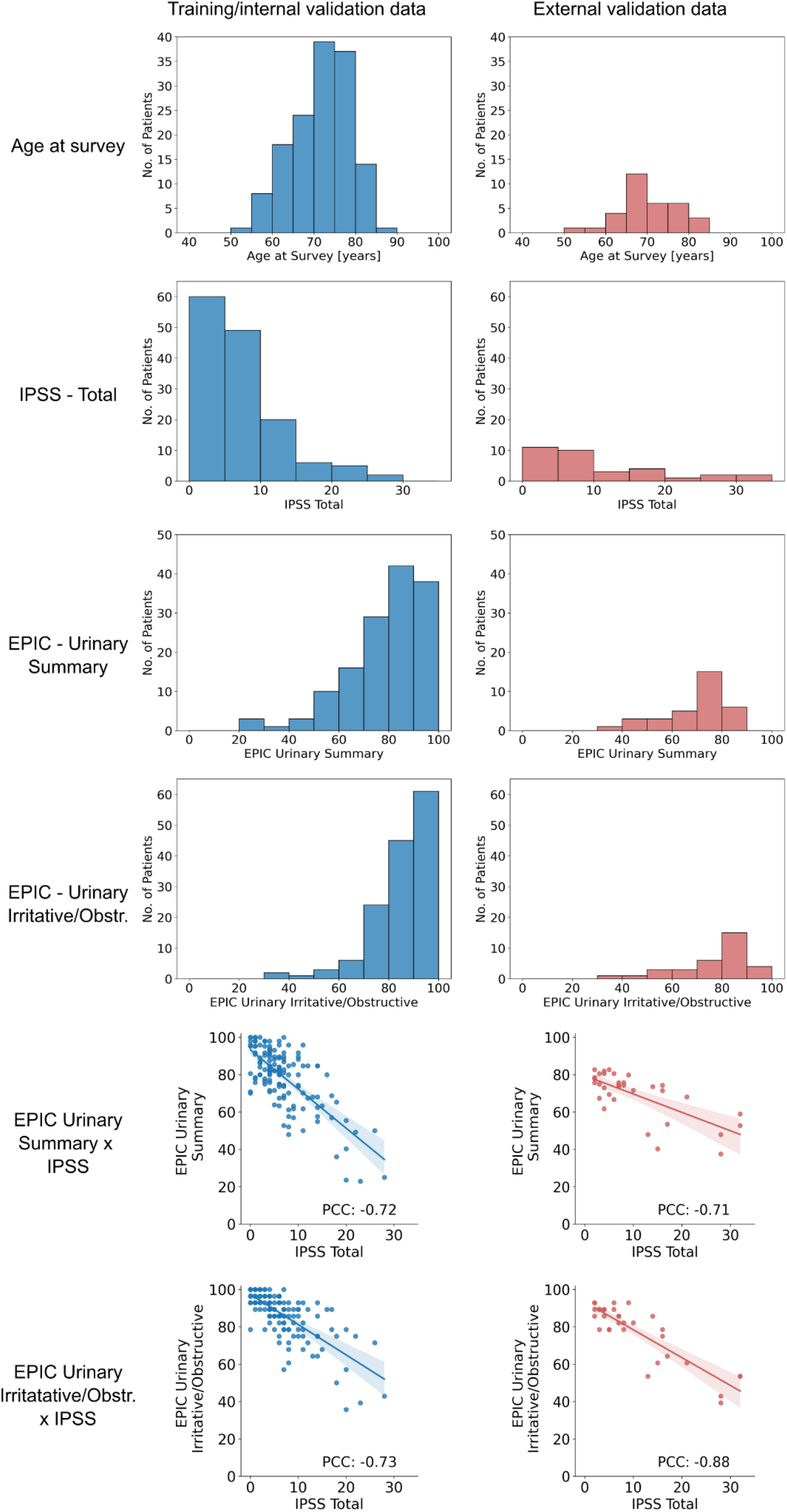
Visualization of selected patient characteristics. Translucent bands in the scatterplots indicate the 95% confidence interval of the regression. *PCC = Pearson Correlation Coefficient*.

We observed a strong negative correlation between the IPSS and both the EPIC Urinary Summary and Irritative/Obstructive subscales with absolute Pearson Correlation Coefficients (PCCs) between 0.71 - 0.88.

The performance of the baseline models is depicted in Table 1 and Figure 2. When using the IPSS as an input, predicting the EPIC Urinary Irritative/Obstructive subscale was more accurate than predicting the EPIC Urinary Summary with mean absolute errors on the external validation set of 6.12 and 10.45, respectively. Conversely, predicting the IPSS was more accurate when using the EPIC Urinary Irritative/Obstructive subscale as an input compared to using the EPIC Urinary Summary with mean absolute errors on the external validation set of 3.88 and 5.13, respectively.

**Table 1.** Mean performance of the baseline models during cross-validation as well as performance during external validation. *CV = Cross-validation*.

| Baseline Models |  |  |  |  |  |  |
| --- | --- | --- | --- | --- | --- | --- |
| Target variable | Input variable | Architecture | Best mean absolute error during CV | Mean absolute error on external data | Min absolute error on external data | Max absolute error on external data |
| EPIC Urinary Summary | IPSS total | Linear regression | 9.19 | 10.45 | 0.09 | 32.75 |
| EPIC Urinary Irritative/Obstructive | IPSS total | Linear regression | 6.41 | 6.12 | 0.37 | 22.78 |
| IPSS total | EPIC Urinary Summary | Linear regression | 3.05 | 5.13 | 0.05 | 20.27 |
| IPSS total | EPIC Urinary Irritative/Obstructive | Linear regression | 2.78 | 3.88 | 0.03 | 14.39 |

**Figure 2.**
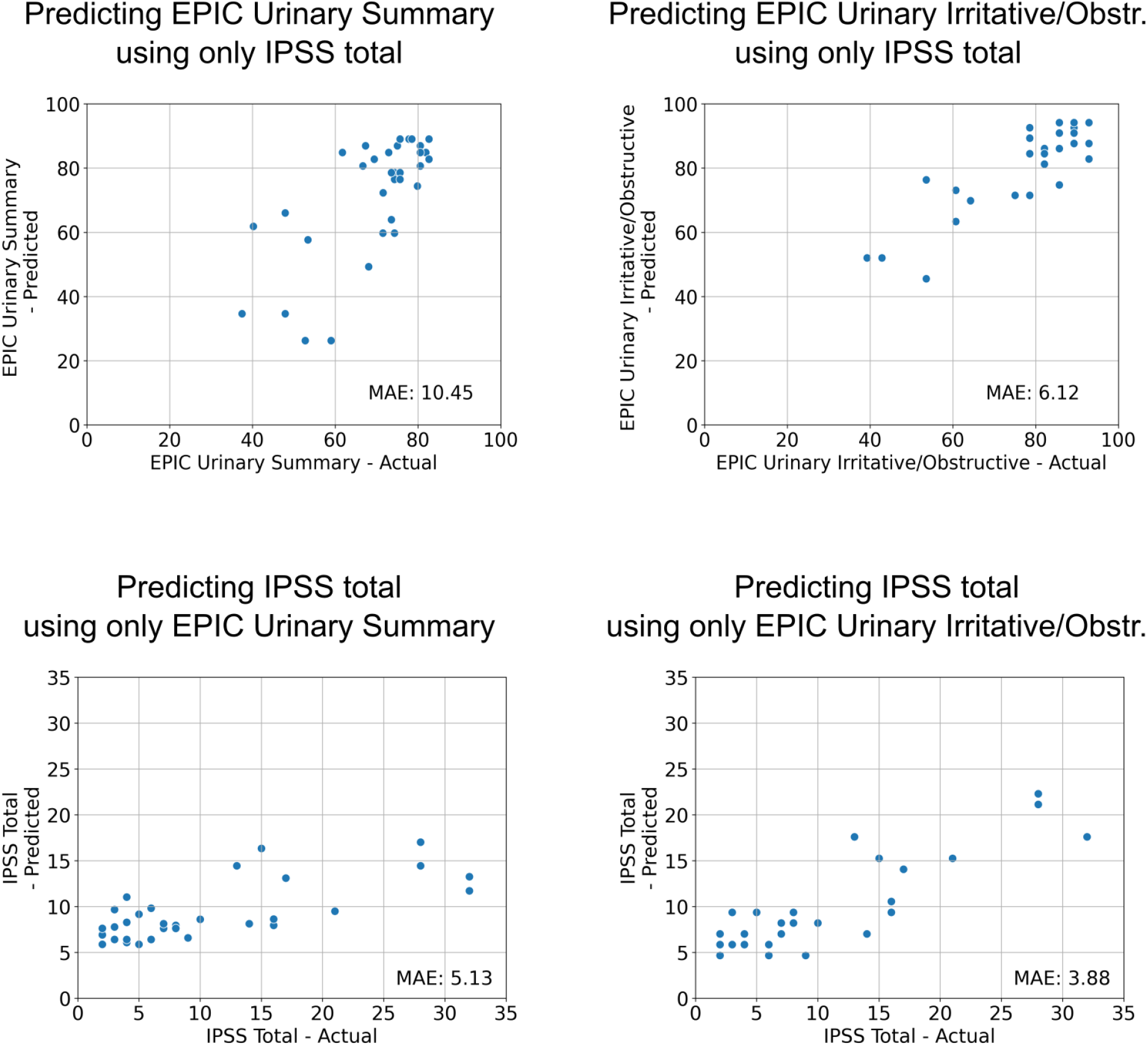
Results of the baseline linear regression models on the external validation set. The coordinates of the dots are determined by the value the model predicted for a given patient in a given questionnaire vs. the actual value the patient obtained. Please note that for the IPSS, lower MAEs are expected due to the scale only ranging from 0-35 compared to the EPIC, which ranges from 0-100. *MAE = Mean Absolute Error*.

The following equations were obtained:

EPIC Urinary Summary = 93.23 - 2.09 * IPSS

EPIC Urinary Irritative/Obstructive = 97.45 - 1.62 * IPSS

IPSS = 26.26 - 0.25 * EPIC Urinary Summary

IPSS = 35.23 - 0.33 * EPIC Urinary Irritative/Obstructive

The performance of the advanced models is depicted in Table 2, Figure 3, and Figure 4. Using all IPSS questions as separate inputs with different model architectures instead of the total score as a single input resulted in only a minor performance improvement when predicting the EPIC Urinary Summary. The mean absolute error of the best advanced model on the external validation set was 9.29 compared to 10.45 for the corresponding baseline model.

**Table 2.**
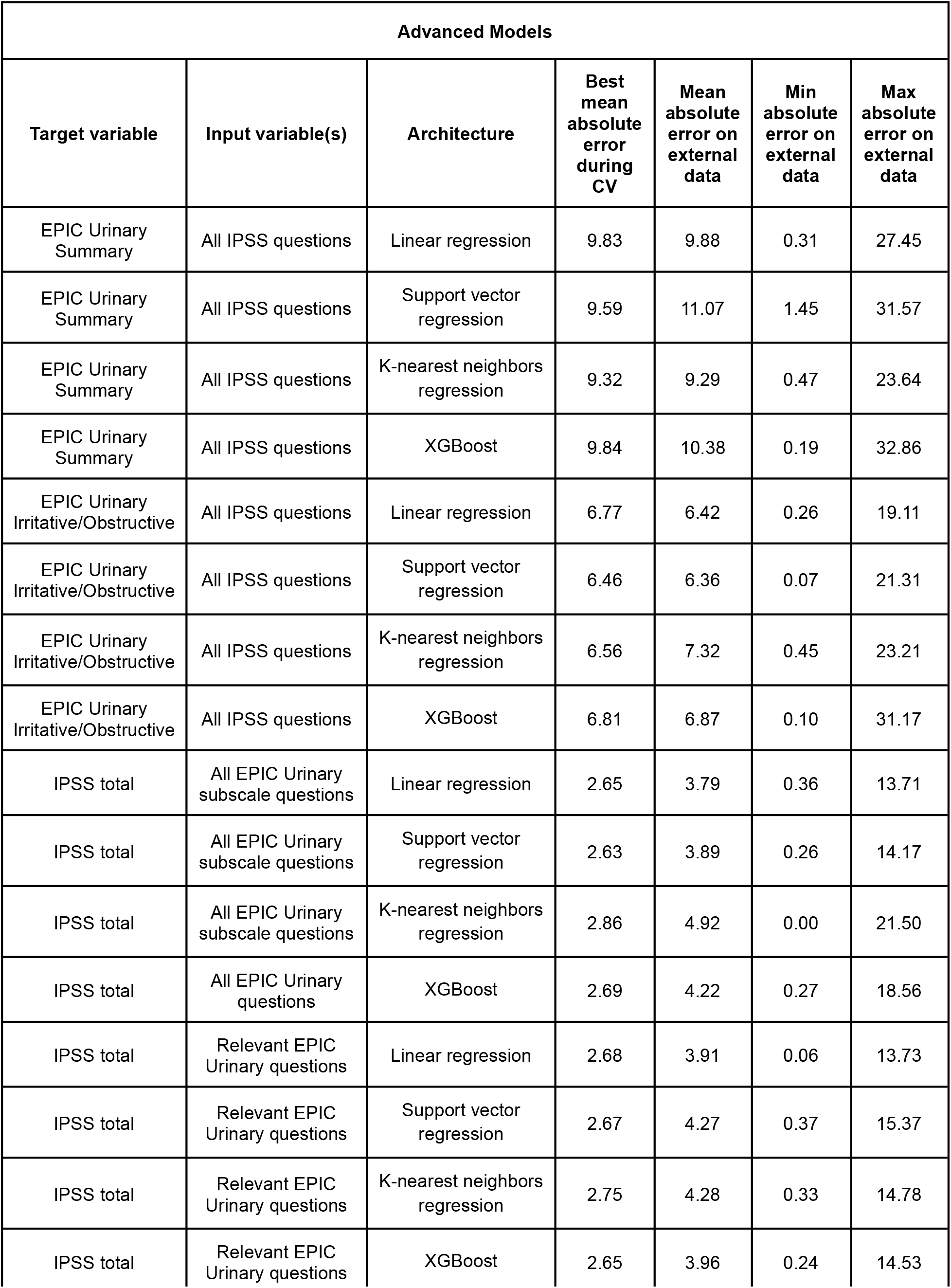
Mean performance of the advanced models during cross-validation as well as performance during external validation. *CV = Cross-validation*.

**Figure 3.**
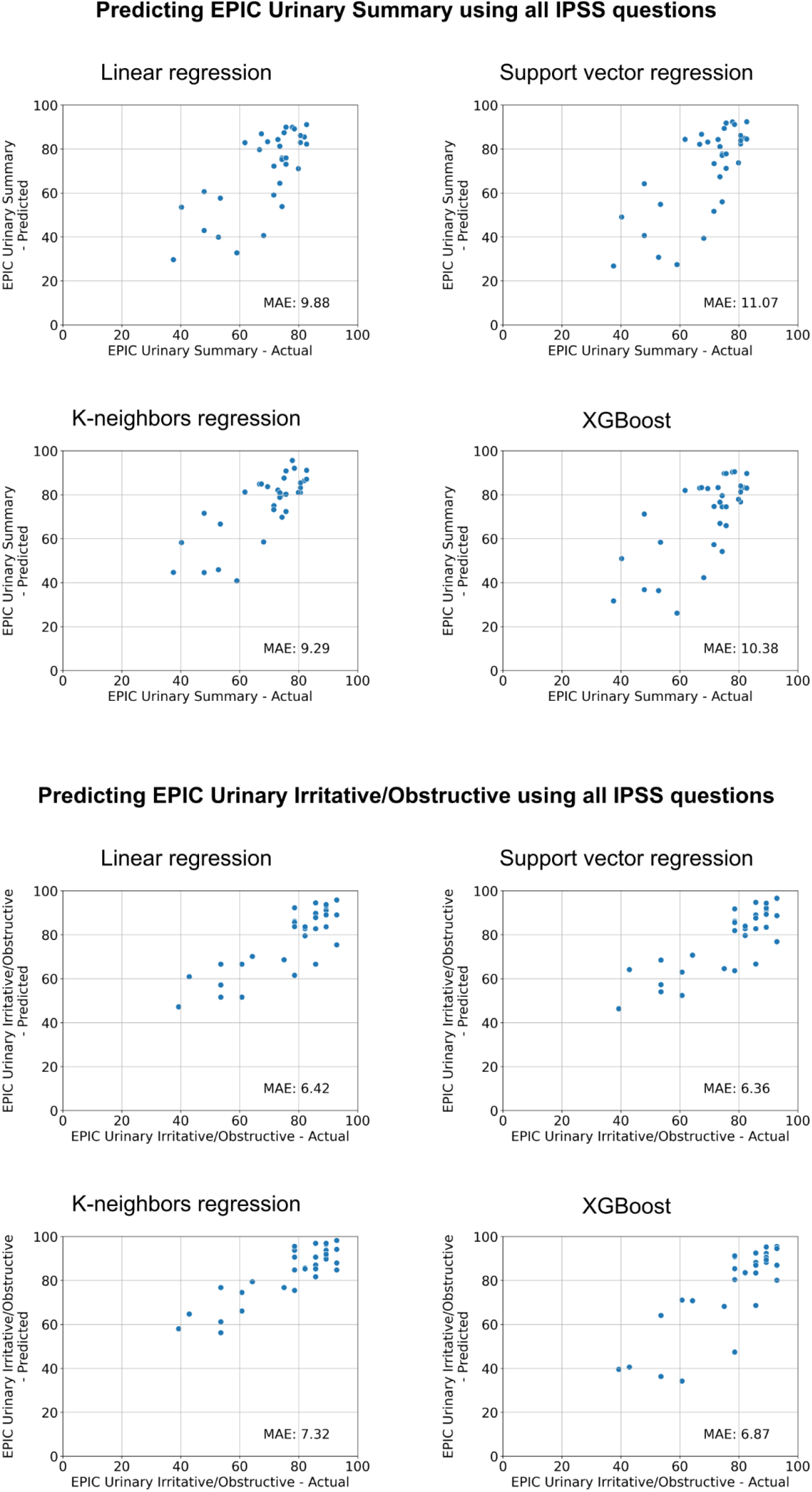
Results of different model architectures for predicting the EPIC Urinary Summary (top) or Urinary Irritative/Obstructive (bottom) subscales on the external validation set using all IPSS questions. *MAE = Mean Absolute Error*.

**Figure 4.**
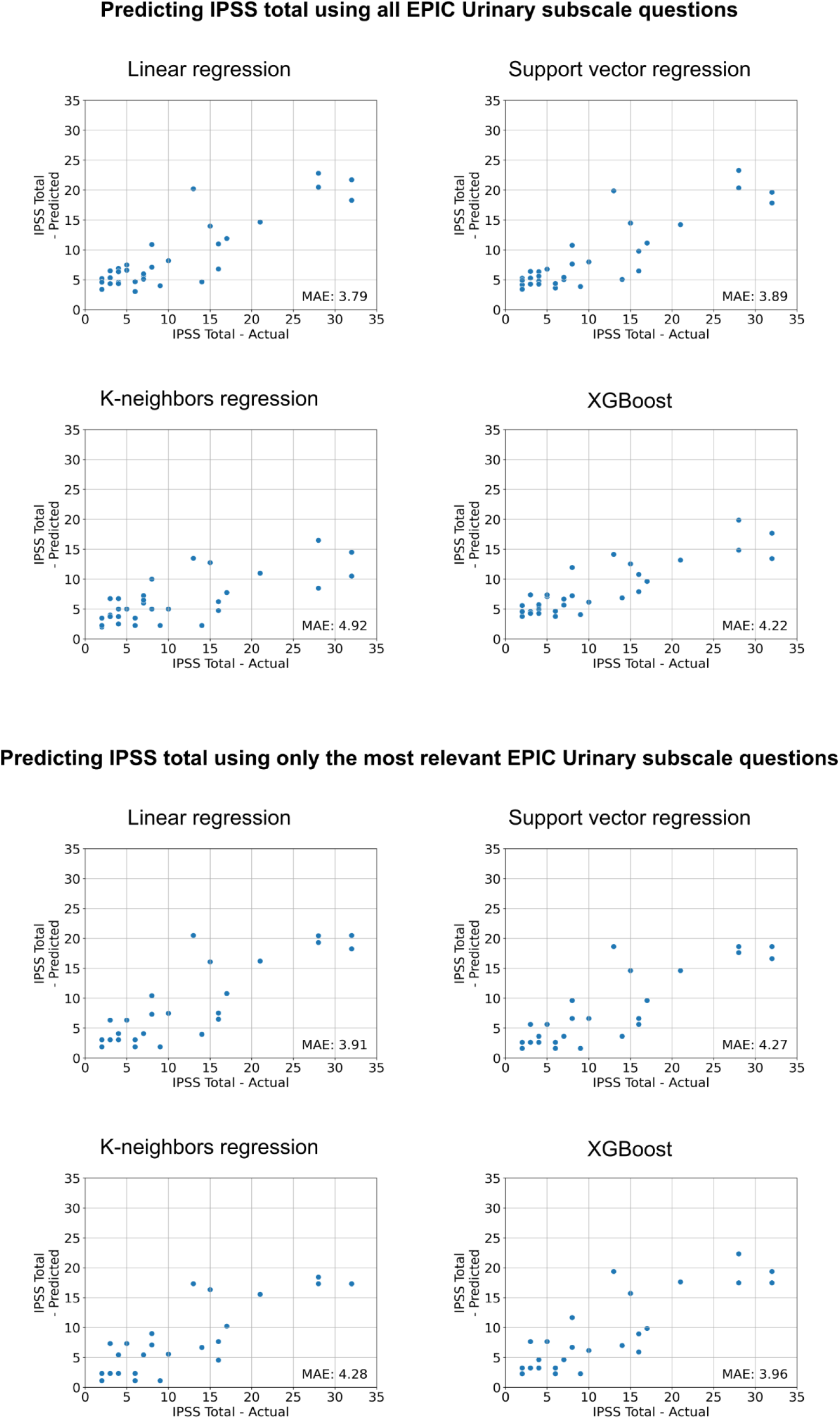
Results of different model architectures for predicting the total IPSS using all EPIC Urinary subscale questions (top) or only the most relevant EPIC Urinary subscale questions (bottom). *MAE = Mean Absolute Error*.

For predicting the EPIC Urinary Irritative/Obstructive subscale, using all IPSS questions did not result in improved performance. The mean absolute error of the best advanced model on the external validation set was 6.36 compared to 6.12 for the corresponding baseline model. Using all EPIC Urinary subscale questions with different model architectures resulted in a mean absolute error of 3.79 on the external validation set, which was an improvement over using only the EPIC Urinary Summary (MAE = 5.13) but only a very minor improvement over using only the EPIC Urinary Irritative/Obstructive subscale (MAE = 3.88)

Using only relevant questions did not result in improved performance.

## Discussion

Our study shows that predicting the IPSS from the EPIC is feasible, especially if the raw questions or the EPIC Urinary Irritative/Obstructive subscale is used as an input. Trying to predict the IPSS using the EPIC Urinary Summary is less accurate, which was to be expected considering that other factors beyond obstructive symptoms influence the Urinary Summary. Blanker and colleagues established a minimal important difference (MID) of 5.2 (95% CI 3.9 to 6.4) for the IPSS in a Dutch cohort.^14^ Both of our baseline models’ mean absolute errors on the external validation set were below that threshold, with an MAE of 5.13 for the model that used the EPIC Urinary Summary as an input and an MAE of 3.88 for the model that used the EPIC Urinary Irritative/Obstructive subscale. However, it should be noted that the maximum absolute errors for a single patient in the external validation set were 20.27 and 14.39, respectively (Table 1). Therefore, while we believe that our equations can be used to compare trial populations, they should not be used to guide clinical decision-making in individual patients. Also, it should be noted that an older publication by Barry et al. suggested a lower MID of 3.1.

Conversely, predicting the EPIC Urinary Irritative/Obstructive subscale using the IPSS was more accurate than predicting the EPIC Urinary Summary with MAEs of 6.12 and 10.45, respectively. Umbehr and colleagues have suggested an MID of 10 for the urinary domain of the EPIC using its German version.^15^ Here again, while the MAEs were at or well below this level, the maximum absolute error in single patients in the external validation set was higher. The fact that more sophisticated model architectures did not result in a significantly improved performance seems reasonable considering the already high degree of correlation between the scores that could very well be modeled using a linear regression.

We have included the equations obtained by the baseline models in an online converter for other researchers to use at https://www.epic-ipss-converter.com/.

In addition to comparing results across studies with different questionnaires, the models could also be used for quality assurance in studies where patients have completed both questionnaires at the same time point: If the result of one questionnaire deviates a lot from the value that was predicted based on the response to the other questionnaire, this might warrant further investigation.

The strengths of this study include the dedicated collection of QoL data, the fact that patients completed both questionnaires at the same point in time, and rigorous preprocessing, which means that patients with missing values were dropped instead of relying on imputations.

Also, an external validation set from another institution in another country than the training cohort was used, which would have highlighted issues with overfitting. Furthermore, a broad range of EPIC and IPSS values was present in both the training and the external validation set.

Limitations of this study include the fact that the German versions of the questionnaires were used and that results for other languages might differ. However, at least for the English versions, this concern is mitigated by the validation processes that the German translations underwent.^7,15^ In addition, we would have preferred to conduct additional validations in previously published studies that used both questionnaires but did not find a study with patient-level data uploaded to a public repository.

Lastly, we believe that improvements in model performance could be achieved by using training data that spans the full range of scores on both questionnaires. While our cohort already represented a variety of scores, no person in the training set had an IPSS greater than 28 or an EPIC Urinary Summary lower than 22, which might have limited the performance of the model in people with very low prostate-related QoL.

## Conclusions

Linear regressions can be used to convert between the IPSS and the EPIC Urinary subscales. More complex model architectures and using the raw answers to the questions did not provide a meaningful performance benefit. While the results of this study can be used to compare results across clinical trials, they should not be used to inform clinical decision-making in individual patients.

